# Analytical Sensitivity of the SalivaDirect™ Assay on the Liberty16 for detecting SARS-CoV-2 B.1.1.529 (“Omicron”)

**DOI:** 10.1101/2022.02.16.22271096

**Authors:** Yen Pei Tan, Mila Al-Halbouni, Ching-Huan Chen, David B. Hirst, Paul J. Pickering

## Abstract

The newly emerged Omicron variant of SARS-CoV-2 has numerous mutations that are not found in other variants of concern (VOCs). Despite acquiring extended functions in adapting to the host-cell environment, the viral genetic variation exerts a potential negative impact on a molecular test, which in turn, compromises public health and safety. The Liberty16 has been clinically validated as a flexible and accessible device system for running the affordable SalivaDirect™ real time PCR detection assay for SARS-CoV-2 especially in low resource settings. Preliminary, based on in-silico sequence analysis, we found that Omicron’s mutation at position 28,311 overlaps with the CDC 2019-nCoV_N1 probe binding region. In order to verify the performance of CDC 2019-nCoV-N1 primers-probe set in detecting the Omicron variant of SARS-CoV-2, plasmids containing Wuhan/WH01/2019 (wild-type) and B.1.1.529 (Omicron) sequences were serially diluted and subsequently directed for SalivaDirect™ RT-qPCR detection on Liberty16 using commercially procured reagents. Our findings provide analytical support for reports that the mutations in the Omicron variant have little or no impact on SalivaDirect™ assay in terms of amplification efficiency and detection sensitivity using either standard and the recently reported fast Liberty16 SalivaDirect™ thermal cycling protocols.

## Introduction

The first known confirmed B.1.1.529 infection was from a specimen collected on 9 November 2021. Based on the evidence presented indicative of a detrimental change in COVID-19 epidemiology and the rapid rise of infections in South Africa, World Health Organization (WHO) has immediately designated B.1.1.529 as a variant of concern (VOC), named Omicron (Figure 1A) (WHO, 2021). Since then, Omicron has spread rapidly through the world and caused the global case count to exceed 1 million daily cases (Singhal, 2022; Taylor, 2022).

**Figure 1.**
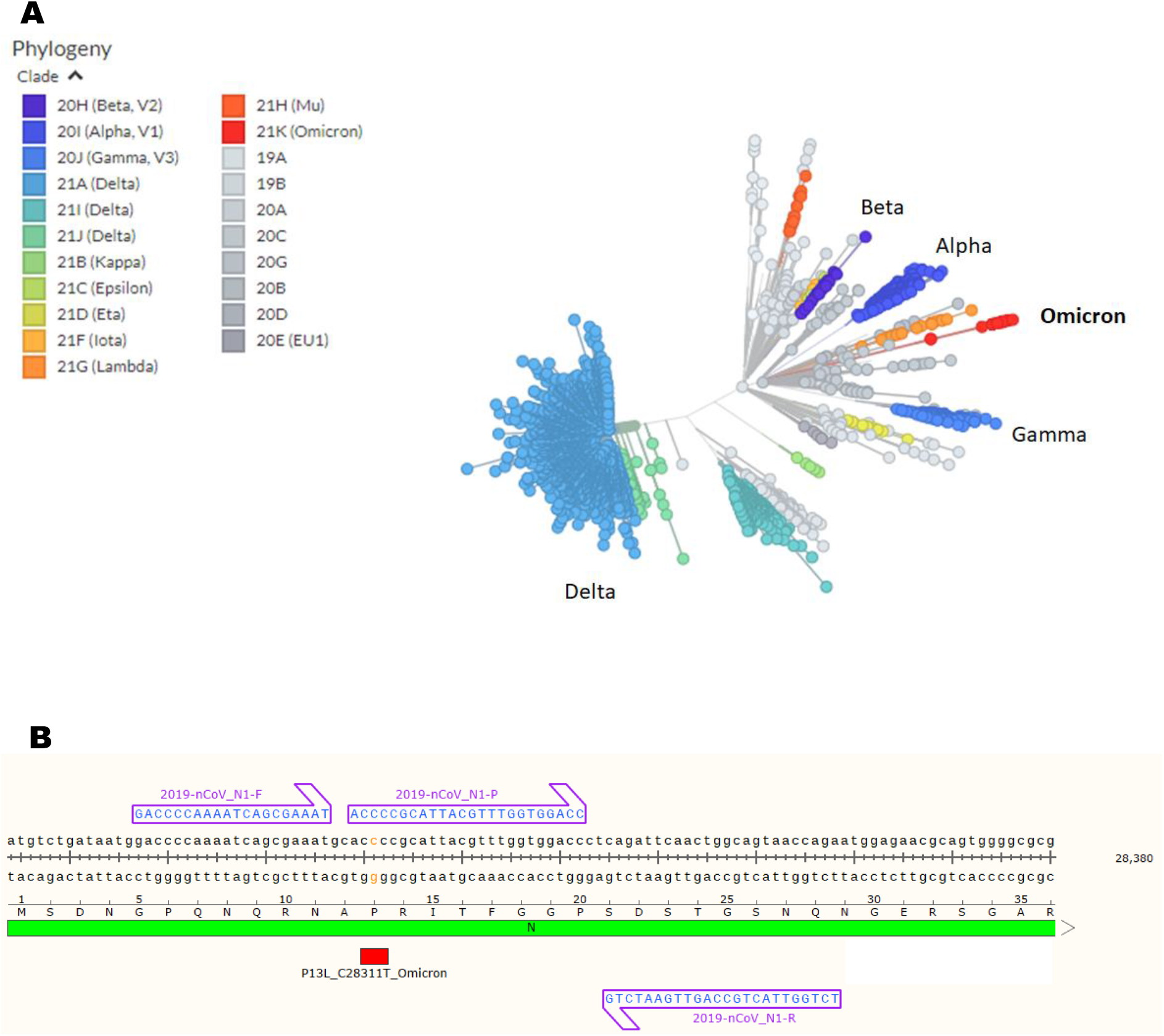
The emergence of Omicron variant and its particular mutation site in 2019-nCoV-N1 probe binding region. A) The Omicron variant and other major or previous variants of concern of SARS-CoV-2 depicted in a tree scaled radially by genetic distance, derived from Nextstrain on 7 December 2021; B) Partial N gene sequences of SAR-CoV-2 isolate Wuhan/WH01/2019 and the binding regions of N1 primers and probe. The mutation site, amino acid P13L or nucleotide base C28311T, of Omicron variant is highlighted in red.

The Omicron variant possesses a large number of new mutations which have not been observed in other strains (Table 1) (Hadfield et al., 2018). Most worrying, over two-thirds of these mutations were found on its spike protein. Like other coronaviruses, the spike protein of SARS-CoV-2 is well recognized as a key component to initiate binding with the host-cell angiotensin-converting enzyme 2 (ACE2) and establish an infection (Zhou et al., 2020). Changes on the Omicron’s spike protein are particularly relevant to increase its affinity for ACE2 receptor and impact host immunity recognition (Cameroni et al., 2021; Redd et al., 2021), though the detailed mechanisms regarding viral pathogenicity and transmissibility remain largely unclear.

**Table 1.** The mutation sites of Omicron variant.

| Gene | Mutations |
| --- | --- |
| ORF1a | K856R, V1887I, S2083-, L2084I, A2710T, T3255I, P3395H, L3674-, S3675-, G3676-, I3758V |
| ORF1b | P314L, I1566V |
| S | A67V, H69-, V70-, T95I, G142-, V143-, Y144-, Y145D, N211-, L212I, G339D, S371L, S373P, S375F, K417N, N440K, G446S, S477N, T478K, E484A, Q493R, G496S, Q498R, N501Y, Y505H, T547K, D614G, H655Y, N679K, P681H, N764K, D796Y, N856K, Q954H, N969K, L981F |
| E | T9I |
| M | D3G, Q19E, A63T |
| N | P13L, E31-, R32-, S33-, R203K, G204R |
| ORF9b | P10S, E27-, N28-, A29- |

Apart from that, the presence of genetic variation in the Omicron variant exerts a potential negative impact on molecular tests which were designed based on the origin sequence. For example, Omicron is highly associated with S-gene target failure (SGTF) on the Taqpath RT-PCR test (Torjesen, 2021). Also, according to FDA’s analysis to date, a single target test developed by Meridian Bioscience, Inc. is already known to fail to detect the SARS-CoV-2 Omicron variant. We found that Omicron’s mutation at position 28,311 overlaps with the CDC 2019-nCoV_N1 probe binding region (Figure 1B). Viral detection using the SalivaDirect™ protocol is solely reliant on the N1 primer-probe (Vogels et al., 2021), and thus, a mutation in this region is potentially impacting on the assay’s sensitivity.

In the presented study, we aimed to assess the impact of the C28311T mutation on analytical PCR performance with the Liberty16 device using the SalivaDirect™ protocol and to verify the assay’s sensitivity in detecting the Omicron variant.

## Materials and Methods

### Assay’s Sensitivity in Detecting SARS-CoV-2 Wuhan/WH01/2019 and B.1.1.529

The partial N gene sequences of SARS-CoV-2 Wuhan/WH01/2019 (wild-type) and B.1.1.529 (Omicron) were generated and cloned into pUC57 at EcoRV/BamHI site (Genscript Biotech Corporation). These plasmids were used to prepare serially diluted analytes for subsequent RT-qPCR detection accordingly to a single-plex version of the SalivaDirect™ protocol run on the Liberty16 System (Yolda-Carr et al., 2022) by using FAM-labelled CDC 2019-nCoV-N1 primers-probe set [Integrated DNA Technologies 2019-nCoV RUO Kit (10006713)] and New England Biolabs Luna Probe One-Step RT-qPCR Mix with UDG (M3019). Additionally, both the standard (Vogels et al., 2021) (95°C, 10s; 55°C, 30s) and fast cycling protocols (Yolda-Carr et al., 2022) (95°C, 2s; 55°C, 5s) were tested.

### Robustness Test

The limit of detection (LoD) for the Liberty16 system was previously confirmed at 12 copies/μL (Yolda-Carr et al., 2022). In the presented study, 2x LoD (24 copies/μL) of plasmid DNA were used for a robustness study. Detection sensitivity was evaluated with 20 replicates per condition.

### Statistical Analysis

The statistical analysis was conducted with a statistics computer software, GraphPad Prism 9.3.1. Differences between the standard and fast cycling protocols were assessed by testing slope differences and Bland-Altman plot.

## Results

### The CDC 2019-nCoV-N1 primers-probe set can detect the Omicron N1 target despite the C28311T mutation

Plasmids containing Wuhan/WH01/2019 (wild-type) and B.1.1.529 (Omicron) sequences were serially diluted into 6-log series, from approximately 5×10^5^ to 5 copies/μL, and subsequently directed for SalivaDirect™ RT-qPCR detection using commercially procured materials. As shown in Figure 2A, the N1 targets were able to be detected across the 6-log dilution series of the mutant and wild-type plasmid DNA. Comparative analysis of these two amplification plots showed no significant difference between the slopes (*P* = 0.7963).

**Figure 2.**
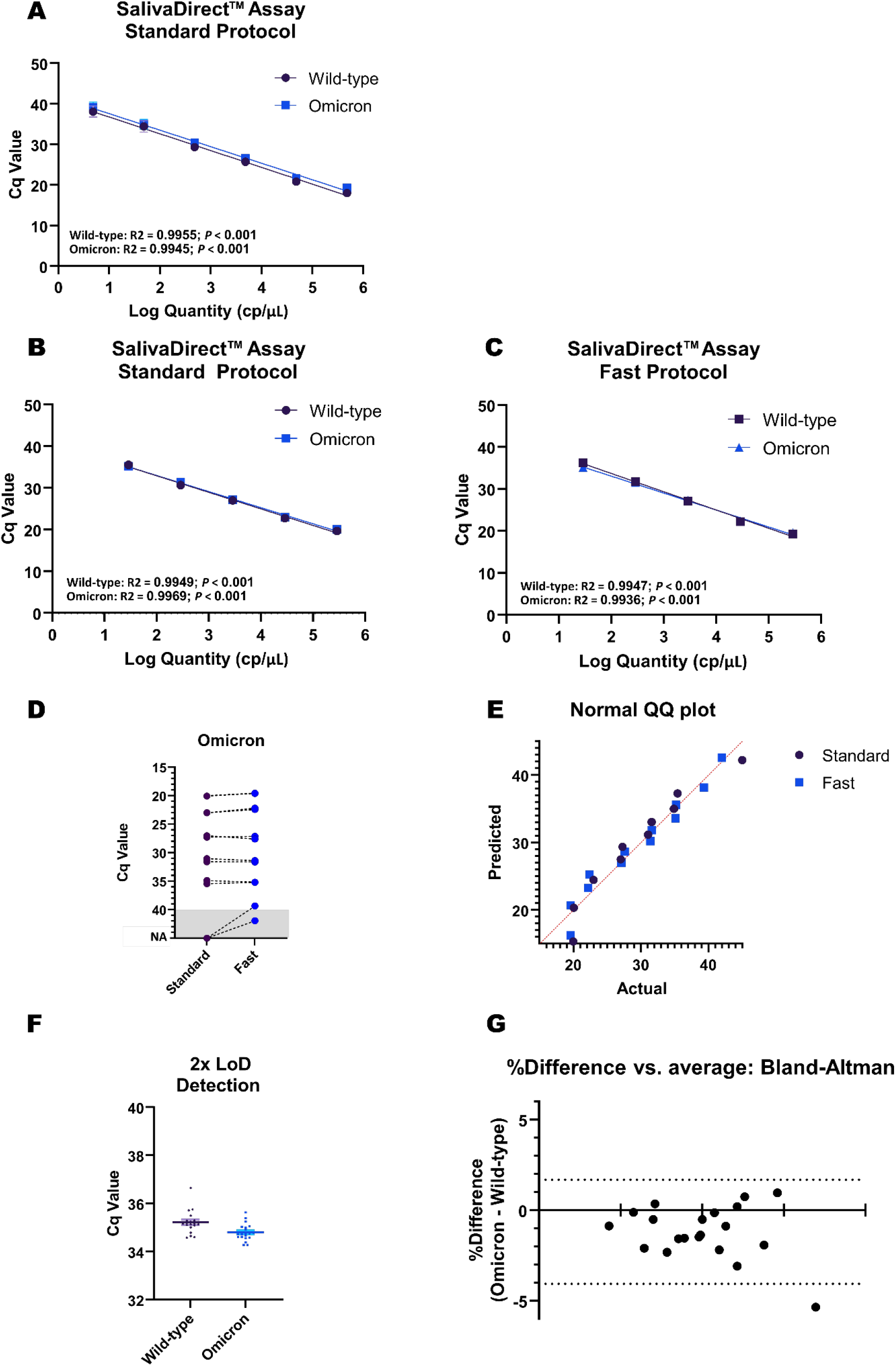
The SalivaDirect™ assay’s sensitivity in detecting wild-type SARS-CoV-2 and Omicron variant. A-B) Linearity performance of SalivaDirect™ assay in detecting wild-type SARS-CoV-2 and Omicron variant using standard cycling protocol; C) Linearity performance of SalivaDirect™ assay in detecting wild-type SARS-CoV-2 and Omicron variant using fast cycling protocol; D) Comparison of the standard SalivaDirect™ RT-qPCR and updated fast cycling protocols on Omicron’s N1 detection; E) QQ plot of the results obtained by two different cycling protocols; F) Robustness test on 2x LoD (n = 20); G) Bland-Altman plot: The agreement between wild-type and Omicron detection; Solid horizontal line shows the mean difference, dotted line shows the 95 % confidence interval of the mean difference.

### Omicron’s N gene can be efficiently detected by using either SalivaDirect™ standard or fast cycling protocols on Liberty16

Next, to assess the agreement between SalivaDirect™ standard protocol and the most recent updated fast cycling protocol on Liberty16, a preliminary study using duplicate samples across a 5-log dilution series (from approximately 3×10^5^ to 30 copies/μL) and a considerably low DNA template concentration (∼3 copies/μL) was conducted. The same samples were run in parallel on Liberty16 devices with different thermal cycling protocols. As expected, the N1 targets were able to be detected across 5-log dilution series of the mutant and wild-type plasmid DNA using either of the two protocols with no significant difference in amplification (Figure 2B and 2C). Remarkably, comparison analysis showed no significant difference between these two protocols in detecting Omicron’s N1 gene (Figure 2D). The data points plotted in the quantile-quantile plot (QQ plot) showed approximately on a straight line, indicating a high positive correlation (Figure 2E). And statistically, high correlation coefficient (R^2^ = 0.9458) was found between these two protocols in detecting either wild-type or Omicron variant of SARS-CoV-2 (with *P* < 0.0001). Hence, we concluded that both protocols worked comparably well in detecting the Omicron variant sequence of SARS-CoV-2.

### The LoD of Omicron detection is comparable to SARS-CoV-2 first isolate Wuhan/WH01/2019

The C28311T mutation did not affect the SalivaDirect™ assay’s sensitivity as 20 out of 20 reactions with 2×LoD (24 copies/μL) were detected for both the wild-type and mutant plasmid DNA (Figure 2E). To further assess the agreement, the Bland-Altman plot was used to assess the existence of a significant bias between these two variants (Figure 2F). As shown, 19/20 of the data points are within 95% limits of agreement. Notably, the only disagreement was caused by an outlier data point testing on wild-type plasmid DNA (Figure 2E and 2F).

## Conclusions

Based on the presented qualitative and quantitative analysis, we conclude that the Omicron C28311T mutation has little or no impact on viral detection using SalivaDirect™ assay. Our findings are similar to recent independent studies (Bei et al., 2021). In addition, our data further confirms that the SalivaDirect™ assay works comparably well using either standard or fast thermal cycling protocols in detecting the Omicron variant sequence. As recently reported, the fast protocol takes just under one hour and is expected to increase sample throughput by more than 60% (Yolda-Carr et al., 2022). This analytical sensitivity study supports the use of the Liberty16 device system with SalivaDirect™ to further facilitate timely access to on-demand screening for infectious respiratory diseases especially within low resource settings.

## Data Availability

All data produced in the present study are available upon reasonable request to the authors

## Aknowledgement

We thank Dr Anne Wylie for her review of this manuscript and Anne and the SalivaDirect™ team for their continued support for SARS-CoV-2 accessible testing solutions for low resource settings.

## Competing Interests

The authors are employees of, and received funding from, Ubiquitome Limited, the manufacturer of Liberty 16 described in the paper. The authors have no other relevant affiliations or financial involvement with any organization or entity with a financial interest in or financial conflict with the subject matter or materials discussed in the manuscript apart from those disclosed.

